# Digital healthcare in public libraries 2002-2025: a scoping review protocol

**DOI:** 10.1101/2024.12.12.24318940

**Authors:** Xavier Chan, Paul Flowers, Megan Crawford

## Abstract

**Objective:** The objective of this scoping review is to explore the use of digital healthcare in public libraries within the literature.

**Introduction:** Accelerated by the COVID-19 pandemic, reliance on digital healthcare has grown in the past decade and will continue to play an increasingly important role in health service provision in the future, including treatment and management of health conditions. An increasing reliance on these digital technologies may exacerbate longstanding inequalities in healthcare access and socioeconomic disparities more widely. Public libraries offer an opportunity to bridge this divide to improve access to digital healthcare. A scoping review will be conducted to explore existing digital health technologies used in public libraries to inform future research.

**Inclusion criteria:** Published studies with adolescents and adults, focusing on digital healthcare in public libraries and conducted in any geographical location will be included in this review. Additional limiters include being published in English and conducted and published between 2002 and 2025. Grey literature sources, including theses, posters and preprints will be included in the review, where relevant.

**Methods:** JBI methodology for Scoping Reviews has been employed for this review. Searches will be conducted using Web of Science, SCOPUS, Pubmed, Library Information and Technology Abstracts (LISTA), Library and Information Science Abstracts(LISA), OpenDisserations and advanced Google search. Sources will be screened by two independent reviewers through 1) title and abstract screening and 2) full text screening. Data will then be charted and presented.

## Introduction

The purpose of this protocol is to outline the background and methodology of a scoping review to explore the use of digital health technologies in public libraries.

The World Health Organisation defines digital health as *“the field of knowledge and practice associated with the development and use of digital technologies to improve health”* (World Health Organisation, 2021). Digital healthcare encompasses technologies including e-health, telehealth and telemedicine, digital therapeutics as well as the Internet of Things, artificial intelligence, big data and robotics (World Health Organisation, 2021). While concepts of telemedicine have existed as early as the 1920s (Moore, 1999), the role of technology has had remarkable impacts to the healthcare field in recent years as digital health has transformed the healthcare landscape, providing innovative solutions to challenges in the health system (Abernethy et al., 2022). The COVID-19 pandemic represented a pivotal shift in the use of digital health, most recognisably through the uptake of telemedicine to remotely provide clinical services in primary care and hospital outpatient appoint such as for consultations and disease management (Mann et al., 2020; Ndayishimiye et al., 2023; Weiner et al., 2021). Other branches of digital health, such as digital therapeutics, have also been accelerated by COVID-19, which may contribute to the transformation of healthcare delivery in years to come (Yao et al., 2024).

A challenge in the rising use of digital healthcare is the digital divide-the unequal distribution between those with and without access to digital technologies, resulting in inequality in the information and tools that digital technologies provide (Cullen, 2001; Saeed & Masters, 2021). The increase in use of digital healthcare to provide clinical services has caused inequalities in healthcare access (Yao et al., 2022). Inequalities in access has been attributed to a lack of network infrastructure, such as in rural areas where broadband reach is limited (DeGuzman et al., 2020). Affordability to both internet access and connected devices also remains a key barrier to internet access leaving socioeconomically vulnerable groups offline (Ofcom, 2022; Yao et al., 2022). Such barriers to accessing digital technology have resulted in ethnically minoritised groups, older adults, individuals with lower socio-economic status, lower levels of education and rural households being more likely to be digitally excluded (Badr et al., 2024). Further, those living with health conditions which impair their ability to physically or psychologically engage with digital technologies are also excluded from digital healthcare (Yao et al., 2022). Other factors influencing access to digital health include digital literacy, where the digital skills required to engage with digital healthcare have been found to be generally higher than those required for everyday use (Badr et al., 2024; Kaihlanen et al., 2022). Finally, attitudes towards digital health technologies may also reinforce the digital divide in healthcare, with poorer experiences of digital health and greater levels of scepticism being associated with lower levels of education and language barriers (Badr et al., 2024). The digital divide results in the exclusion of vulnerable groups from accessing digital healthcare, which exacerbates socioeconomic, health and digital inequalities (Sun et al., 2020). It is therefore critical to consider avenues and implement strategies to mitigate the effects of digital exclusion in the rise in digital healthcare.

Public libraries are uniquely positioned within the community to support the increase in access to digital healthcare and could serve as hubs for patrons to access digital healthcare. Public libraries have previously been utilised as places for health for underserved populations (Elia, 2019; Rubenstein, 2018). Through partnerships with community organisations, academic institutions and local governments, public libraries have been able to increase access to health services, through linkage and direct access to healthcare (Philbin et al., 2018). The COVID-19 pandemic necessitated the rapid adaptation of public libraries to support communities in novel ways, including through the provision of health information (Ma et al., 2023), and supporting wellbeing virtually, such as hosting online yoga and partnering with health care exchange agencies to facilitate uptake of health insurance (Acosta et al., 2023; Ashiq et al., 2022). Further, owing to the widespread availability of high-speed internet and free public access to computers, public libraries operate as gateways to accessing digital technologies for individuals who do not have access to the internet within their homes, which represented around a quarter of the library users in a survey conducted in the UK (Leguina et al., 2021). Public libraries not only provide access to the internet and connected devices, but also support the use of the technology, for example, to use the internet for information about health (Baker & Sugarman, 2023). Given the roles public libraries have played in supporting health, and providing access to digital technologies for underserved populations, public libraries could be considered ideal locations within communities to support access to digital healthcare.

Despite the potential, the role of public libraries to improve access to digital healthcare remains relatively under-explored. Through a search of MEDLINE and Web of Science, only one review was identified using digital health technologies-a narrative review and research agenda specifically assessing the use of telemedicine video visits in public libraries (DeGuzman et al., 2022). This review identified only one publication using telemedicine video visits in public libraries, and public libraries was not the primary focus of the study (DeGuzman et al., 2022; McIlhenny et al., 2011). At present, no systematic or scoping reviews specifically exploring the broader use of digital health in public libraries have been identified. A preliminary search of MEDLINE, the Cochrane Database of Systematic Reviews and *JBI Evidence Synthesis* was conducted and no current or underway systematic reviews or scoping reviews on the topic were found. Given continuous innovation in digital health technologies and the increasing reliance on digital technologies in healthcare accelerated by the COVID-19 pandemic, it is of increasing necessity to explore the use of digital health technologies in public libraries. Hence, this review will inform and may make recommendations for future research and service development.

To identify how future services within public libraries could be developed to improve access to digital healthcare, a scoping review has been identified as an appropriate methodology to map the broader literature on digital healthcare in public libraries. This will be structured according to 1) the health topics addressed by digital healthcare in public libraries, 2) the types of digital healthcare used in public libraries 3) the theoretical approaches to the research, and 4) the barriers and facilitators to the implementation of digital health in public libraries, where applicable. A scoping review was selected over a systematic review due to the more inclusive approach offered by a scoping review, whilst still maintaining transparency and replicability (Munn et al., 2018). Overall, the aim of this scoping review is to explore the literature studying digital healthcare provided in public libraries.

### Review questions

RQ1: What health topics are addressed in studies on digital healthcare within public libraries, as reported in the literature?

RQ2: What types of digital healthcare have been used in public libraries within the literature?

RQ3: Which, if any, theories/frameworks have been used in research on digital healthcare in public libraries?

RQ4: How many sources refer to barriers and facilitators to implementing digital healthcare in public libraries in the literature?

### Inclusion/Exclusion criteria

#### Population

##### Inclusion

Studies involving populations of adults and adolescents will be included. For adolescents, terminology will include but will not be limited to “teenager”, “teen” “adolescent”, “youth”, “young people”. Studies with subpopulation analysis (e.g. age, socioeconomic status, ethnicity) will also be included only where the findings regarding public libraries are extractable and independent of other contexts.

##### Exclusion

Studies involving child populations (including but not limited to “child”) or with participants <10 years of age.

##### Rationale

While the targets of subsequent studies of this review are primarily adults, adolescents (considered 10-19 by WHO (World Health Organisation, 2024)) have been considered in this review due to limited number of publications from preliminary searches focusing on adult samples only.

#### Concept

##### Inclusion

Studies focusing on any patient/public facing digital health in public libraries. Where studies include public libraries in their methodology, but is not the primary focus of the study, the publication will be included only where the findings regarding public libraries is extractable.

##### Exclusion

- Studies relating to digital healthcare not in public library settings-this includes limited-access, institutional or private libraries (e.g. university or school), and other community settings (e.g. GP or community centre).
- Studies relating to non-digital healthcare within public libraries
- Studies where public libraries are included but are not the primary focus of the study, and where the findings are not extractable

##### Rationale

The focus of the review is specifically to explore the literature studying digital healthcare in public libraries. An inclusive approach to the concept has been adopted due to the limited number of studies exclusively using implementation science to assess the provision of digital healthcare in public libraries. However, regarding setting, institutional/non-public libraries, and other community settings have been excluded from the review since public libraries are uniquely involved in supporting health (e.g. health information) and the provision of access to and support of the use of digital technology.

#### Context

##### Inclusion

- Studies based in any geographical location will be included.
- The data within the studies must have been collected during or after 2002.

##### Exclusion

- No publications will be excluded according to geographical location.
- Publications where data has been collected prior to 2002 will be excluded from this review.

##### Rationale

- Given the exploratory nature of this review, at present there is no reason to exclusively explore a specific geographical region.
- 2002 reflects that implementation of the People’s Network (Library and Information Commission, 1997)-the introduction of free internet and computer access in public libraries in the UK.

#### Sources

##### Inclusion

Research published in peer reviewed journals with primary data collection-quantitative, qualitative or mixed methods. Grey literature will also be considered (Doctoral theses, posters, pre-prints).

##### Exclusion

Reviews, opinion articles, blogs, protocols, conference abstracts

##### Rationale

Given the exploratory nature of this scoping review and limited numbers of publications found in preliminary searches, some grey literature sources will be included to broaden the scope of the search strategy and to reduce the effect of publication bias.

#### Publication language

##### Inclusion

Only publications written in English language will be included in this review.

##### Exclusion

Publications that are written in any language other than English will be excluded.

##### Rationale

Only publications written in English language will be used in this review due to limited resources for translation.

#### Publication year

##### Inclusion

Research published between 2002 and 2025

##### Exclusion

Research published prior to 2002

##### Rationale

(see context)

## Methods

This review will be conducted in line with JBI methodology for scoping reviews (Peters et al., 2020). Preferred Reporting Items for Systematic reviews and Meta-Analyses extension for Scoping Reviews (PRISMA-ScR) checklist (Tricco et al., 2018) will be used to detailing the results of the full review.

### Quality Appraisal

In line with JBI guidelines for conducting a scoping review (Peters et al., 2020), the quality of the available literature will not be assessed for this review.

### Search strategy

A university librarian was consulted with during the development of the research methodology for this scoping review. Preliminary searches through Web of Science Core Collection using the terms “digital health” and “public library” were conducted to develop the inclusion/exclusion criteria in October 2024.

In line with JBI guidelines, a three-step search strategy has been developed for this scoping review. An initial limited search of Web of Science and SCOPUS has been conducted to identify salient terms contained in the titles and abstracts of relevant articles. These have been used, along with the index terms used to describe the articles, to develop a preliminary search strategy. The search strategy is subject to development and will be adapted for each database, accounting for differences in syntax and terminology (example search strategy can be seen in appendix 1). Once finalised, the search strategy will be employed for a second search across 6 databases-Web of Science Core Collection, SCOPUS, Library, Information Science and Technology Abstracts (LISTA) and Library and Information Science Abstracts (LISA) and PubMed and OpenDissertations for Doctoral theses. Grey literature searches will also be conducted using advanced Google searches to identify relevant grey literature sources not available in the included databases, such as pre-prints and conference posters. Where available, references from the searches will be screened to identify other relevant sources not captured by the search strategy. Further, where appropriate, grey literature sources, such as conference abstracts, will be used to identify sources that would be captured in this review, such as published or pre-published articles.

### Source selection

Identified sources from conducted searches will be uploaded to COVIDENCE (Veritas Health Innovation, 2024) and undergo automatic deduplication, and a manual deduplication screening. Following a trial screening to ensure reliability between three independent reviewers, the first round of screening will be conducted using titles and abstracts to assess inclusion/exclusion according to the criteria outlined in the PCC framework, with each citation being screened by two reviewers (XC will screen all articles, and two MSc students will share the task of double screening).

Once title and abstract screening has been completed, the full text of each citation will be screened in detail according to the inclusion/exclusion criteria by two independent reviewers (XC will screen all articles, and two MSc students will share the task of double screening). The reason for exclusion of citations at full text-screening will be reported in the review. Should there be discrepancies in inclusion or exclusion of citations by reviewers at any point throughout the screening process, these will be discussed on a case-by-case basis, employing a third reviewer (MC) where consensus cannot be made.

### Data charting

Data will be extracted from papers included in the scoping review following full-text screening using a data extraction table using COVIDENCE, adapted by the author from the JBI manual for evidence (Peters et al., 2020). This charting table (available in appendix 2) will detail information relating to the citation (author, year), study (e.g. aims, sample size, research methods, data collection, analysis), population (e.g. characteristics, age, demographics), concept (focus of intervention, implementation theories employed, relevant findings, barriers and facilitators to implementation) and context (country, year data collected). Where this data has not been made available, authors will be contacted to request the missing data if appropriate. The extraction chart is subject to modification throughout the extraction process and a description of these changes will be outlined within the final scoping review.

### Data analysis and presentation

Frequency analysis of details of the publications and studies will be used, and presented in tables to outline the use of digital healthcare in public libraries in the literature. The research questions will then each be addressed by synthesis of the literature into a table, followed by a narrative summary of the findings. For research question 4, where applicable, the barriers and facilitators to implementation will be extracted verbatim from each source and interpreted by the author using standardised behavioural statements. The barriers and facilitators will be synthesised where the barriers/facilitators are similar. The Theoretical Domains Framework (Cane et al., 2012) will be employed to systematically categorise and code the theoretical underpinnings of these barriers and facilitators to implementing digital health interventions in public libraries. Subsequently the behaviour change wheel (BCW; Michie et al., 2011) will be used to identify intervention functions and behaviour change techniques (BCTs) that could be useful to enhance future implementation of digital healthcare in public libraries using behaviour change technique taxonomy version 1 (BCTTv1).

## Data Availability

All data produced in the present study are available upon reasonable request to the authors.

## Acknowledgements

This review has been conducted as part of a PhD project supervised by Dr Megan Crawford and Professor Paul Flowers (University of Strathclyde).

## Funding

This scoping review is funded by a PhD studentship granted to the first author (XC) at the University of Strathclyde by the Scottish Graduate School of Social Sciences (SGSSS), which is jointly funded by the Economic and Social Research Council (ESRC) and the Scottish Funding Council. The funding was acquired by Dr Megan Crawford and Professor Paul Flowers.

## Author contributions

**Xavier Chan**: Conceptualisation, writing and editing, methodology

**Prof. Paul Flowers**: Conceptualisation, supervision, reviewing and editing, funding acquisition

**Dr Megan Crawford**: Conceptualisation, supervision, reviewing and editing, funding acquisition

## Conflicts of interest

There are no conflicts of interest in this project.

## Appendices

**Appendix 1:** Example search strategy.

| Database | Web of Science |
| --- | --- |
| Digital health | ("digital health" OR "e-health" OR "electronic health" OR "m-health" OR "mobile health" OR telehealth OR telemedicine OR "digital medicine" OR "digital therap*" OR teletherap* OR "digital mental health" OR ((digital OR online OR virtual OR "comput*" OR app OR tele* OR technolog* OR remote) AND health AND (care OR intervention OR therap* OR treatment OR "mental health" OR service))) |
| Public Library | ("librar*" OR "public library" OR "community library")<br>NOT ( "Public Library of Science" OR "academic librar*" OR "hospital library" OR "medical library" OR "digital library" OR "online library" OR "health librar*" OR "school library" OR "university library" ) |
| Limiters | 2002-2024<br>Published in English only |

**Appendix 2:** Data extraction tool (adapted from JBI manual)

|  |  |
| --- | --- |
| Citation details | • Author<br>• Title<br>• Publication Year<br>• Journal<br>• Volume<br>• Issue<br>• Page number |
|  | • Source type (e.g. journal article, thesis) |
| Study details | • What are the aims/objectives of study?<br>• Study design<br>• Methods of data collection (e.g. interview, survey)<br>• Qualitative or quantitative data<br>• Data analysis technique(s)<br>• Length of study |
| Participants | • Total number of participants<br>• Eligibility criteria<br>○ Inclusion<br>○ Exclusion<br>• Demographic information<br>○ Age (mean or range)<br>○ Gender<br>○ Ethnicity<br>○ Socioeconomic status<br>○ Education<br>○ Urban/rural population |
|  | ○ Health condition(s)<br>○ English proficiency |
| Concept | • Health topic (e.g. diabetes)<br>• Type of healthcare (e.g. health information, non-clinical or clinical)<br>• Intervention (developed for study in public libraries) or service (existing intervention used in public library)<br>• Digital health used (e.g. digital therapeutic, telemedicine video visit, online health information service)<br>• Theories/Framework used<br>• Barriers and facilitators<br>○ Barriers to implementation<br>○ Facilitators to implementation<br>• Recommendations for implementation |
| Context | • Country<br>• Year data collected |

## Notes

### Competing Interest Statement

The authors have declared no competing interest.

### Summary of Updates

This manuscript has been updated to correct an error in the abstract of the previous version referring to temporal limiters being articles conducted and published between 2002 - 2024. This has been updated to be 2002 - 2025. Further, clarification has been provided within the section titled data charting, where 'barriers and facilitators' have been amended to be 'barriers and facilitators to implementation'. Within the data analysis and presentation section, other corrections regarding language have been made, including expansion of the acronyms BCW (behaviour change wheel), BCT (behaviour change technique) and BCTTV1 (Behaviour change technique taxonomy version 1) for readability. Finally, additional updates to this section clarify that the barriers and facilitators will first be extracted verbatim, then interpreted using standardised behavioural statements. The barriers and facilitators will then be synthesised according to their similarity.

